# A Divide and Conquer Strategy against the Covid-19 Pandemic?!

**DOI:** 10.1101/2020.05.05.20092155

**Authors:** Patrick Mangat

## Abstract

The concern about (socio-)economic consequences of collective lockdowns in the Covid-19 pandemic calls for alternative strategies. We consider a *divide and conquer strategy* in which a high risk group (HRG) is put on strict isolation, whereas the remainder of the population is exposed to the virus, building up immunity against Covid-19. The question is whether this strategy may suppress the effective reproduction number below the critical value of 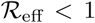 without further lockdown once the HRG is released from isolation. While this proposal appears already rather academic, we show that 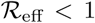 can only be obtained provided that the HRG is less than ~ 20 − 30% of the total population. Hence, this strategy is likely to fail in countries with a HRG larger than the given upper bound. In addition, we argue that the maximum infection rate occurring in this strategy is likely to exceed realistic capacities of most health care systems. While the conclusion is rather negative in this regard, we emphasise that the strategy of *stopping the curve* at an early stage of the Covid-19 pandemic has a chance to work out. The required duration of the lockdown is estimated to be τ ~ 14 days/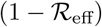 (up to some order one factor) for 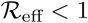, provided a systematic tracing strategy of new infections exists for the subsequent relaxation phase. In this context we also argue why 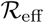 remains the crucial parameter which needs to be accurately monitored and controlled.

## 1 Introduction

Most countries have been locked down for more than six weeks already. The goal of the lockdown strategy is to avoid a collapse of the health care systems. However, this goes at the cost of significant economic decline including growing debts and unemployment (for studies on (socio-)economic impacts see e.g. [1–7]). While the Covid-19 pandemic directly threatens our health, the link between economic growth and our health is rather implicit and non-trivial. The lockdown particularly protects the so-called high risk group (HRG) [8–10], whereas the economic consequences will affect the whole population. Consequently, in Germany (and most likely in other countries as well) doubts have been raised whether the current mitigation measures are proportionate.

In particular, it has been proposed whether it may suffice to impose isolations only for the HRG, while the rest of the population is exposed to the virus (various discussions can be found in German newspapers such as *Spiegel Online*). The hope behind this proposal is that the decline of the economic growth is minimised while the health care system is kept stable (because the non-HRG is less likely to require intensive treatment), see e.g. [1] for related work in this direction. Apart from various practical issues with this proposal, this strategy makes only sense if the effective reproduction number eventually drops below 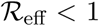 once the HRG can be released from the isolation. Of course, the hope is that 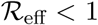 is reached purely due to the immunity of the non-HRG. This would mean that the Covid-19 pandemic is in control even without further measures afterwards.

We call this the *divide and conquer strategy*, because the population is divided into a HRG and a non-HRG. Using the SIR-model [11] we prove that the divide and conquer strategy requires the HRG to be less than approximately 20 – 30% of the whole population. Hence, at least in Germany this strategy will most likely be unsuccessful. In addition, we argue that realistic capacities of the German health care system are likely to be exceeded due to the infections of the non-HRG in this strategy. We use the SIR-model, because it captures the relevant mechanisms of a pandemic despite its simplicity.

This note is organised as follows. We briefly recall the basics of the SIR-model including an application to Covid-19 and the *flatten the curve strategy*. Afterwards we apply the SIR-model to the *divide and conquer strategy*. Finally, we summarise the importance of the reproduction number in this crisis and propose a simple formula to estimate how long one needs to keep 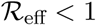 until a lockdown can be relaxed.

## 2 A primer on the SIR-model

This section recalls the basics of the SIR-model (following [12–14]) and briefly demonstrates the dynamics in the Covid-19 pandemic as well as the problem with *flattening the curve* in this context. Readers who are familiar with this model can skip to the next section.

### 2.1 The SIR-model

The SIR-model divides the population into susceptible (*S*), infectious (*I*) and recovered (*R*) people. This is the origin of the name of the model developed in [11].

The total population *N* is then given by

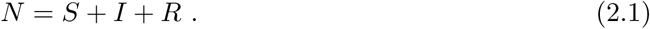

The SIR-model assumes constant population, i.e. no deaths, births and migration. An infectious person will infect a susceptible person at a transmission rate *β*, and will recover at a recovery rate *γ*. Any recovered person is assumed to be immune against the disease forever. The system of differential equations then reads

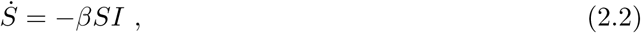

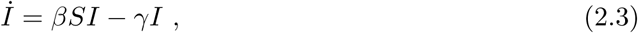

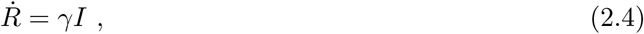

where *Ṡ = dS/dt* (derivative of *S* with respect to time t) and likewise for *İ* and *Ṙ*. By adding up those three differential equations one can easily see that *Ṅ* = 0, i.e. the total number of population is indeed conserved.

A crucial parameter is the *basic reproduction number* 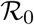, which can be written at *t* = 0 as

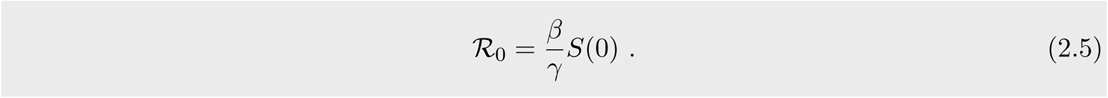

The basic reproduction number tells us to how many people the disease is transmitted on average due to *one* infectious individual. To see where (2.5) comes from, we suppose that at *t* = 0 we have *I*(0) = 1. The probability of this infectious person to remain infected at time *t* is *p*(*t*) = *e*^−^*^γt^*. Hence, the expectation value for the number of people this single person will infect, is given by

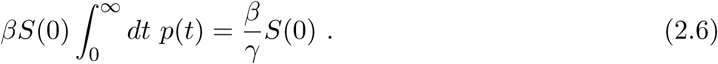

Given 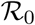, the *effective reproduction number* 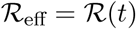 is

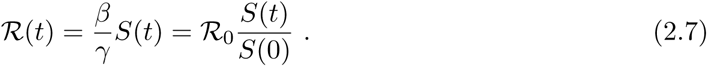

Hence, as the pandemic is progressing, one infectious person will infect fewer and fewer people on average, because more and more people recover in the course of this pandemic.

Therefore, the effective reproduction number decreases automatically and, eventually, it will drop below the critical value of 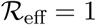. This is when *herd immunity* is reached.

To understand this in detail, we rewrite (2.3) in terms of 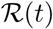:

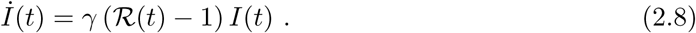

By linearising the differential equations one can rigorously prove that an epidemic occurs if and only if 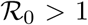. A less rigorous argument is the following. Initially, 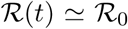 because *S*(*t*) ≃ *S*(0) ≃ *N* initially in a large population with only few infectious and recovered people. Then, (2.8) immediately implies an exponential growth of infectious people, namely

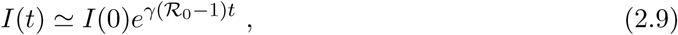

at the *beginning* of the epidemic. For 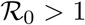 the disease is spreading exponentially, whereas for 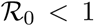 the recovery is faster than the spread leading to exponential suppression. At the critical value of 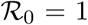 the number of new infections is compensated by the number of recovered people (per time interval).

But once the number of immune people increases to a significant amount, the *I*-curve deviates from an exponential function, and eventually reaches its maximum, at which 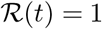. This is when *herd immunity* is reached, because from then on *İ*(*t*) < 0 until *I*(*t*) → 0.

At herd immunity the proportion of people that have been infected is given by

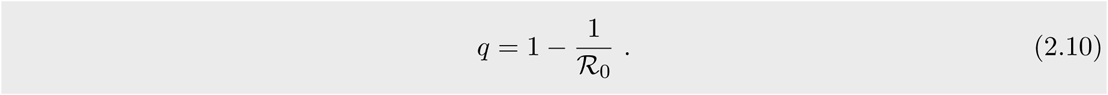

To see this, note that at *t = t_H_* (time of herd immunity) we have

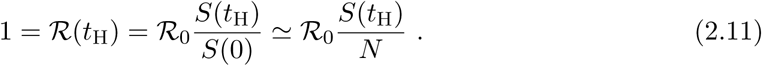

By definition, *q* ≡ 1 − *S*(*t_H_*)/*N* and the assertion (2.10) follows.

Despite herd immunity there will still be further new infections at a rate

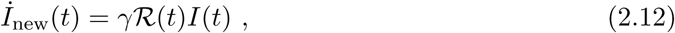

i. e. at *t* ≫ *t_H_* a significantly larger fraction than *q* will have had contact with the disease.

Let

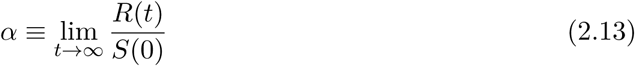

be the fraction of people who will eventually get into contact with the virus in the course of the pandemic (for *S*(0) ≃ *N* this represents the fraction of the whole population). We can rearrange (2.2) and (2.4) to

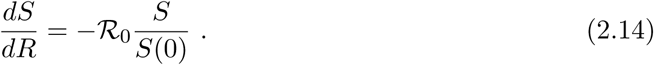

Solving this differential equation results in

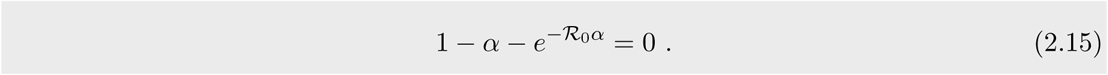

For *α* ≠ 0 this equation has only a solution in terms of the Lambert *W* function. We observe that 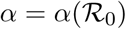, i.e. the quantity *α* is purely determined by the reproduction number. This equation will be important in Section 3.

Another important equation regards the maximum number of people that are simultaneously infected, *I*_max_, reached at herd immunity. Solving *İ*(*t_H_*) = 0 implies *S*(*t_H_*) = *γ/β*. Furthermore, (2.2) and (2.3) imply

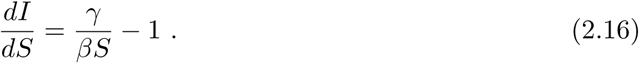

The general solution is

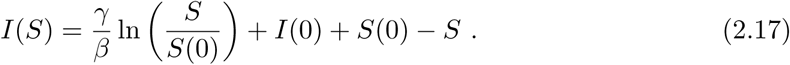

At *t = t_H_* we then have

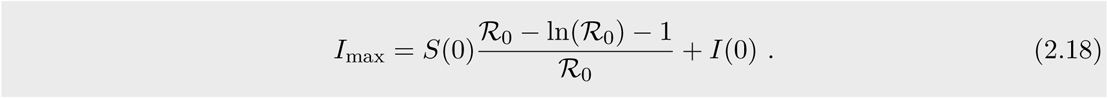

But more important than this number is the maximum of the daily new infections, which should be below the health care capacities. The condition for the maximum of new infections reads

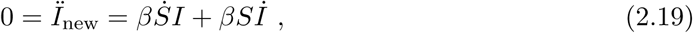

which yields

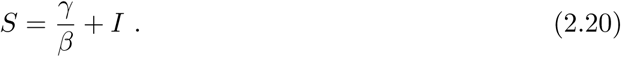

This can be inserted into (2.17), yielding a complicated equation in *I*. But in the limit 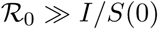 we obtain

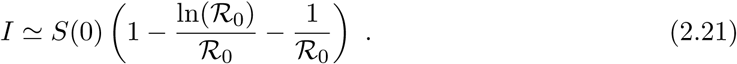

The limit 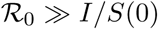 is roughly satisfied for 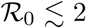. Hence the maximum number of new infections in this limit is

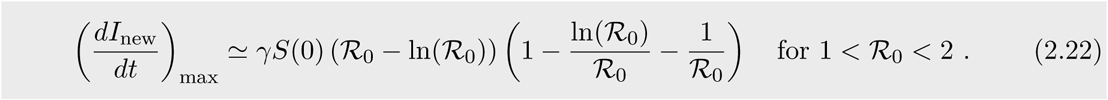

If 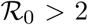 then the result becomes imprecise and, therefore, requires either the inclusion of higher order corrections in 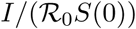 or a numerical approach. This number of new daily infections must be smaller than the capacity of the health care system. (Of course, this capacity is non-trivial to estimate.)

### 2.2 Application to the Covid-19 pandemic

We apply the SIR-model to the Covid-19 pandemic in Germany. The intention is merely to illustrate the underlying dynamics rather than making any predictions. In the past months a lot of effort has been put into mathematical modelling of the Covid-19 pandemic, see e.g. [15–28] for an extensive, yet incomplete, list.

First of all, we notice that the SIR-model is approximately suitable to describe the Covid-19 pandemic, because to present knowledge a recovered person is immune against reinfections for some significant time at least.^1^ However, some infections lead to death. But technically, dead people can be treated like recovered people in the SIR-model. Therefore, we do not need to modify the model for the purpose of this note. In more realistic models of epidemiology one should also include the status “exposed”, in which a person got infected without being infectious yet. This is accounted for in the SEIR-model. Many more complications can be introduced, such as asymptomatic cases etc., see e.g. [16] for a sophisticated model tailored to describe Covid-19 pandemic.

In the SIR-model of Covid-19 we can set the recovery rate *γ* = 1/14 day^−1^ ≃ 0.071 day^−1^, because typically it requires 14 days to recover from Covid-19.

Estimations of 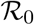 range from 1.4 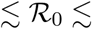 5.7, see [29–32] and references therein. A precise value does not only depend on the virus itself but also on various external conditions such as hygienic standards, people’s behaviour, density of population (see [33]), or possibly even climate (see e.g. [34]). For the first two weeks of March we can estimate for Germany [35]:

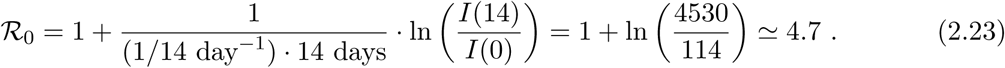

Of course, this estimation is problematic, because neither the number of tests nor the number of undetected cases is known (a transparent approach has been presented in [22]). Since the testing has become more intense in this period, the above is likely to overestimate 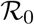. Nevertheless our estimate is still consistent with the figures computed in [16].

In Figure 1 the underlying dynamics of the SIR-model is shown. The eqs. (2.2) to (2.4) have been integrated numerically via the Euler method with initial conditions *S*(0) = 80 · 10^6^, *I*(0) = 114 and *R*(0) = 0. We observe that smaller 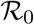 suppresses the peak *I*_max_ (see also (2.18) for an analytical expression), and also shifts the time of herd immunity, *t*_H_, further into the future. In particular, for a Covid-19 pandemic without any mitigation attempts we can estimate 2 months ≲ *t_H_* ≲ 6 months. This estimation intends to give an idea about time scales of a pandemic. Moreover, unsurprisingly, the plateau of the R-curve decreases for smaller 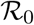. Using (2.15) we obtain *α*(2.0) ≃ 0.80, *α*(3.0) = 0.94 and *α*(4.0) = 0.98. Hence, without any measures between 80% – 98% of the population will have had contact with the virus at the end of the pandemic. Note that those figures are not in contrast to the often quoted 60% – 70% of infections. Those numbers refer to the moment of herd immunity. Indeed, from (2.10) we get *q*(2.0) = 50%, *q*(3.0) = 67% and *q*(4.0) = 75%.

**Figure 1:**
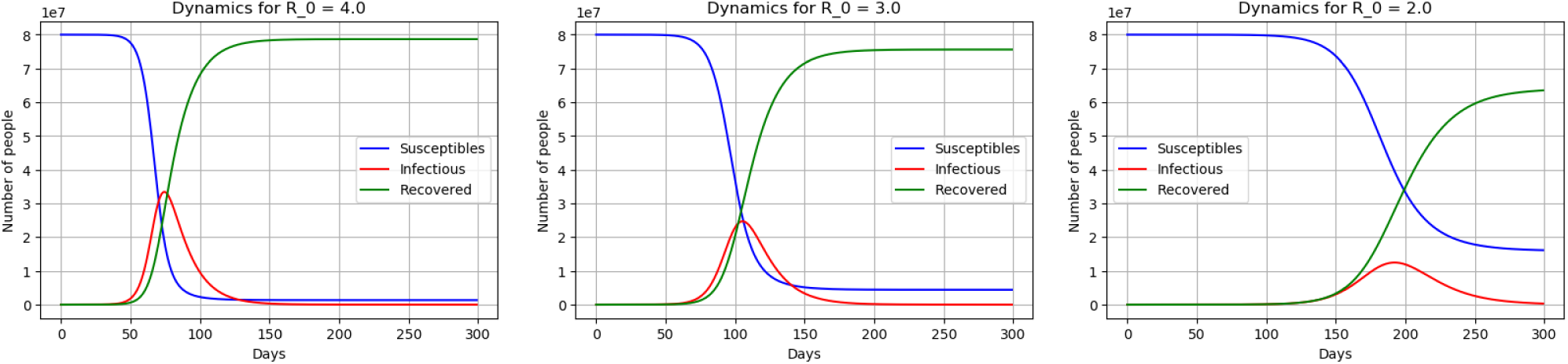
These plots show the dynamics of the Covid-19 pandemic for reproduction numbers 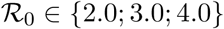. The number of susceptible (blue), infectious (red) and recovered (green) people is plotted against time (in days). It is emphasised that these plots are *not* suitable for predictions. Instead they merely reveal the underlying dynamics of the SIR-model.

Furthermore we observe that even in the case of 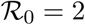 more that 13% of the population will be infectious at the peak of the pandemic. This is clearly unacceptable given the relatively significant case fatality rates. The situation therefore triggered many ideas how to mitigate worst case scenarios within this pandemic (see e.g. [1; 2; 17; 19; 33; 36–45]). Rather intuitive strategies that had been suggested are the concepts of *flattening the curve* (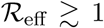) and *stopping the curve* (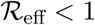).

### 2.3 Flattening the curve and fine-tuning in the Covid-19 pandemic

In this section we point out a problem with flattening the curve. The curve that needs to be flattened is the *I*-curve, or, more strictly speaking, the rate of new infections curve, *İ*_new_(*t*) = *βS*(*t*)*I*(*t*). The latter curve will reach its peak before reaching herd immunity. This curve must be flattened in a way such that the peak will be below some upper bound *C* due to capacities of the health care system. It must be ensured that

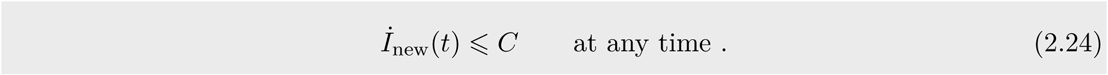

A simplified estimation of *C* may be as follows. Let *n*_bed_ = 30 · 10^3^ be the number of available beds in intensive care units of hospitals to treat severe cases at an average duration of ∆*t* = 10 days. The probability of any random infectious person to require intensive care is assumed to be *p =* 5%. Then,

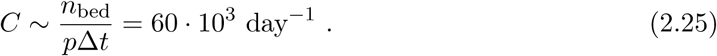

This figure can only be seen as a very rough estimate, because each of its variables is just an estimate. In addition, any constraint regarding health care staff or further necessary surgery equipment has been ignored. Nevertheless, this number of *C* ~ 60 · 10^3^ day^−1^ suffices to point out the problem with the concept of flattening the curve below *C*.

To see this we plot *İ*_new_(*t*) for the parameter range 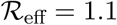 to 1.3. We use the notation 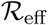 instead of 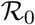 because the rather small numbers are assumed to arise from some measures (like social distancing). The plot can be easily obtained numerically by computing *I(t* + 1) − *I*(*t*). The result is shown in Figure 2. We chose initial conditions as of 2020-05-03 without aiming at guessing further unknown cases. The results shown are not meant to be a prediction of the future.

**Figure 2:**
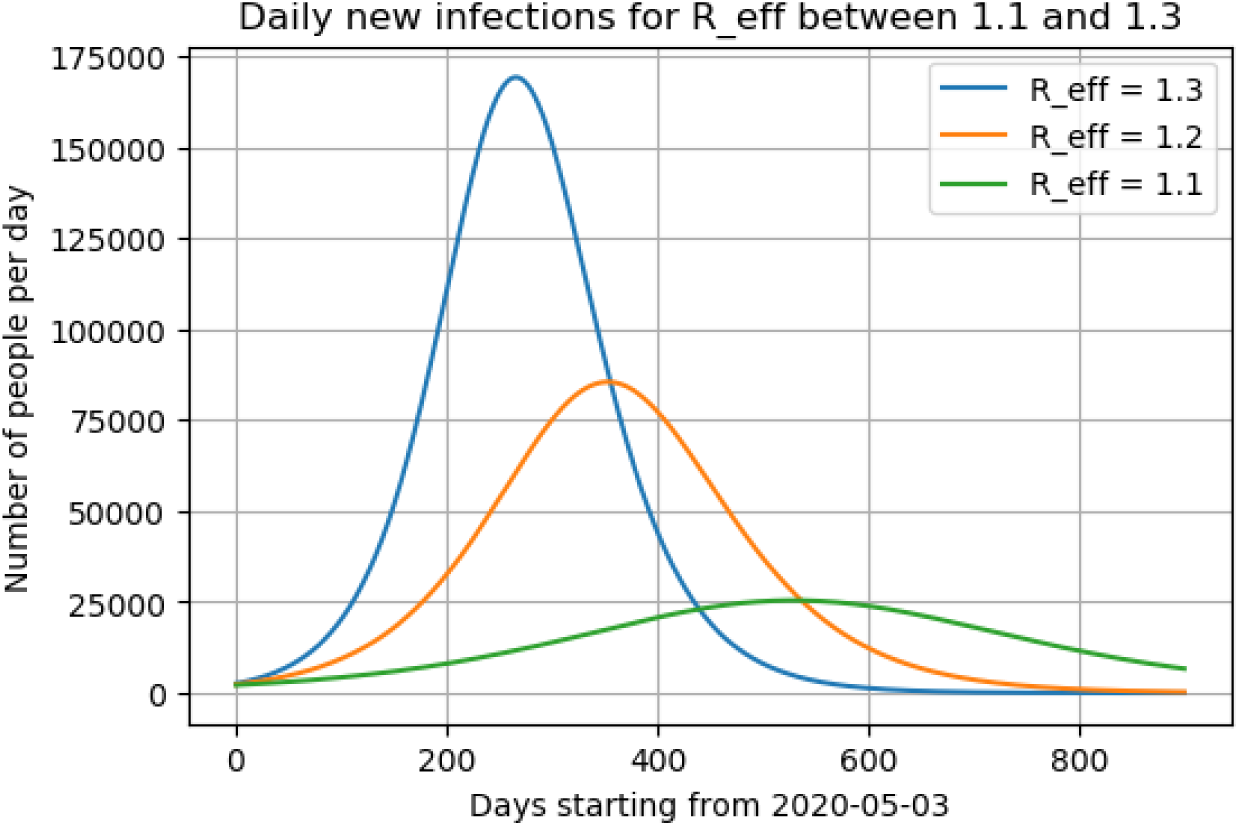
This plot shows the expected daily infections for the Covid-19 pandemic in Germany for 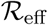 between 1.1 and 1.3. The initial conditions were set by the publicly available data for 2020-05-03 [35].

We find that already the first decimal place of 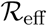 has strong influence on the maximum height of the curve *İ_new_* (see (2.22) for an approximative formula). A jump from 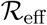 from 1.1 to 1.3 suffices to increase the peak of daily new infections by a factor larger than 6. In particular we see that 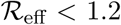 is needed to remain below the bound of 60 · 10^3^ infections per day. Therefore, the parameter range of the effective reproduction number in *flattening the curve* is 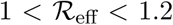, and hence very tiny. We call this requirement *fine-tuning*. The weaker the health care system the stronger the fine-tuning needs to be. Given the error bars in measuring 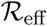 (see [16]) it seems quite challenging to keep control of the pandemic within this strategy. In particular, the precise impact of different measures on 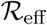 is unknown at present. Furthermore, the strategy *flatten the curve* seems to require a duration of larger than one year! Consequently, *flattening the curve* seems not be the way to go in the Covid-19 pandemic.

## 3 The divide and conquer strategy

Since the measures against the Covid-19 pandemic particularly protect the high risk group (HRG) the following *divide and conquer strategy* may appear worth considering: Only the HRG is strictly isolated while no significant measures are imposed on the non-HRG. We assume that infectious people only exist among the non-HRG. This situation is illustrated on the left hand side of Figure 3. Due to the dynamics explained in Section 2 most of the initially susceptible people become immune. If the proportion of immune people is large enough it is conceivable that 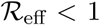 even upon releasing the HRG from the isolation (depicted on the right hand side of Figure 3).

**Figure 3:**
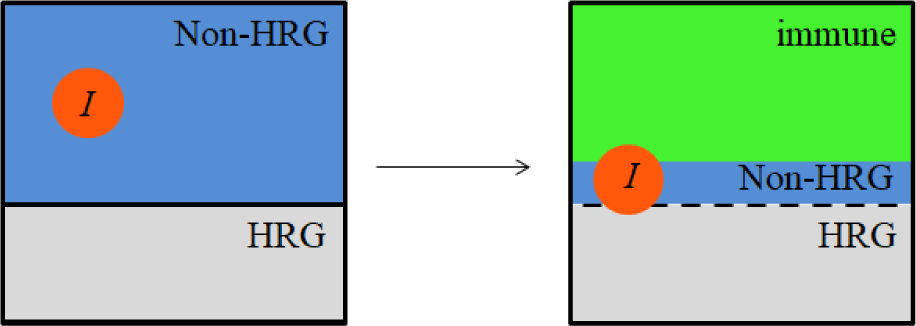
Illustration of the divide and conquer scenario. The whole population is initially split into a high risk group (HRG) and a non-HRG. Infectious individuals are assumed to exist only among the non-HRG, while the HRG is strictly isolated. As time evolves most of the non-HRG become immune. The HRG is released from the isolation if 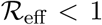 can be ensured due to the large fraction of immune people.

One may hope that such a strategy can keep the economy alive without risking too many human lives.

The question we want to answer is: If we isolate *Q* people (from *N* people in total) how large can we make the fraction *Q/N* to ensure 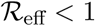 after the isolation simply due to herd immunity?

To answer this question we distinguish between two cases. In the first case, the *dynamical divide and conquer strategy*, we assume that all infections occur merely due to the dynamics of the SIR-model without any further intervention. In the second case, the *divide and conquer strategy with intentional infections*, we admit interventions to maximise immunity.

In what follows we are ignorant about all practical problems with these strategies such as strictly isolating millions of people for many months or even more. In this sense this discussion remains rather academic.

### 3.1 Dynamical divide and conquer strategy

Since we isolate *Q* people initially, we have *S*(0) ≃ *N − Q* susceptible people left (assuming *I*(0) ≪ *N*). The initial effective reproduction number corresponding to the left picture in Figure 3 is

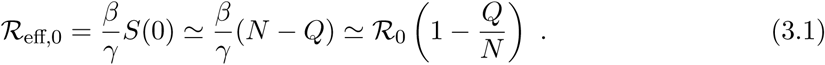

Now, one might suggest to isolate sufficiently many people to reach 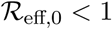 in the first place. But then the isolation would have to last until effective vaccination is available. Therefore, the divide and conquer strategy relies on herd immunity *after* releasing the *Q* people from isolation.

If we wait long enough for the susceptible population to get infected and to recover, we will have approximately

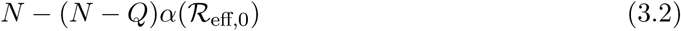

susceptible people on the right hand side of Figure 3. Recall that 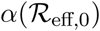 is a solution to (2.15).

Hence, the new effective reproduction number becomes

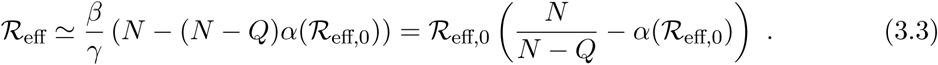

By imposing 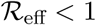 we obtain the condition

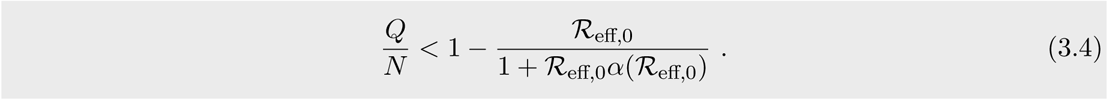

The optimum can be quantified by solving (2.15) numerically. We find that

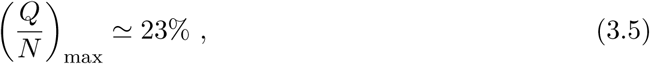

which is reached for 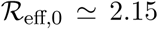. The plot of (*Q/N*) as a function of 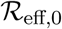 is shown in Figure 4.

**Figure 4:**
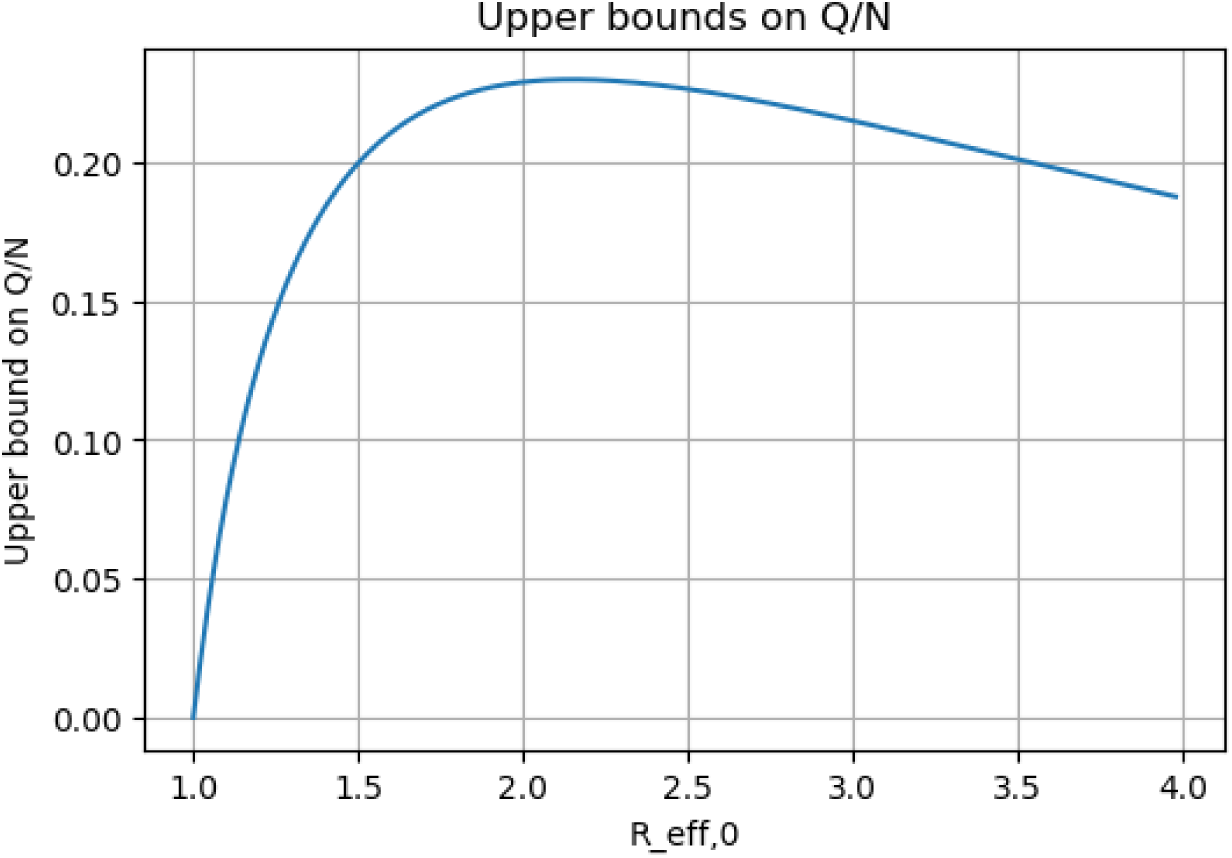
Upper bounds of *Q/N* as a function of 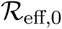

This result is a problem, because the HRG is certainly larger than 23%. For instance, the fraction of people older than 60 years is in Germany above 25%, wo are included in the HRG if we follow the definition of [9]. Note that in [8] and [10] people are counted to the HRG if they are older than 65 and 70 years, respectively. However, also pregnant women, smokers, chronically ill or obese people, etc. count to the HRG as well. An investigation into estimating the size of the HRG in Germany is beyond the scope of this note, especially because a precise definition of the criteria of the HRG is still under discussion among experts.

Another problem is the peak in the curve of new daily infections among the non-HRG. At 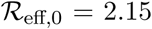 one obtains 9.51 · 10^5^ infections per day (reached after 5 to 6 months from the starting point of divide and conquer). Hence, even if the probability for an individual of the non-HRG to require intensive treatment is ten times smaller than assumed in Section 2.3 the capacities of the health care system are exceeded. To avoid a collapse one rather needs 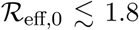 (leading to a maximum of 5.8 · 10^5^ infections per day). This, however, reduces the maximum of *Q/N* by only 1%. In general, from Figure 4 it becomes clear that the *divide and conquer strategy* needs to accept large numbers of new daily infections (see also [1]).

### 3.2 Divide and conquer strategy with intentional infections

There is a loophole that invalidates (3.4) if we admit intentional infections. Thereby one could artificially increase the number of immune people close to *N − Q*. Then,

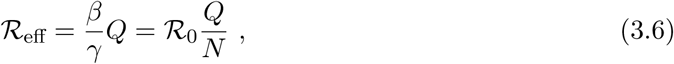

or, requiring 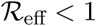,

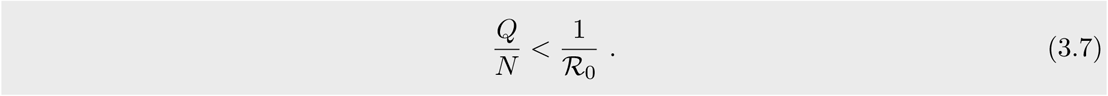

Note that in this inequality 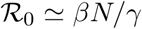 is the basic reproduction number for the SARS-CoV-2 virus without any particular measures. The result is not surprising given the formula (2.10). Therefore, assuming 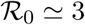, we get

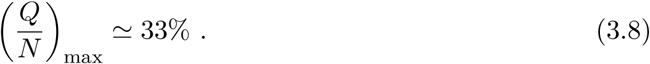

This may still turn out to be too small to accommodate the whole HRG among the isolated people.

In any case, even if we assume that we could isolate 50% of the population (the HRG in the first place), while we ensure that the other 50% become immune, we lower 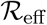 down to 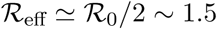 at the end of isolation. This is the rate at which the initially isolated people will get infected. As demonstrated in Section 2.3 this implies a collapse of the health care system, especially because the HRG is more likely to end up in an intensive care unit. Hence, further restrictions would still be unavoidable. Therefore, the *divide and conquer strategy* seems to provide no reasonable solution.

### 3.3 Equipartition strategy?

Finally we address some interesting effect that occurs upon dividing the population into *n* equal, non-interacting, groups (see [17]).

First, suppose that the population is split into *n* pairwise isolated groups such that

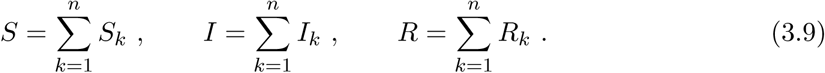

Each group individually obeys the differential equations of the SIR-model:

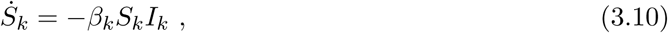

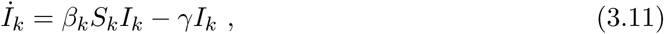

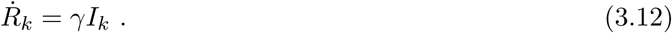

We assume that only the recovery rate *γ* is equal among all groups.

Let us now assume an equipartition, i.e. *S_k_ = S/n, I_k_ = I/n* and *R_k_ = R/n*. The first differential equation then becomes

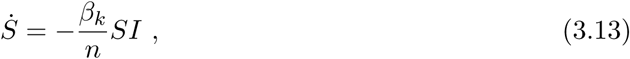

and the overall transmission rate is reduced to *β = β_k_/n* relative to each group (this observation was made in [17] in a more general model). Unfortunately, this effect is undone in the reproduction rates. We have

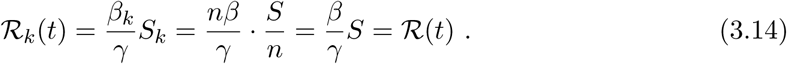

Since it is the reproduction number that determines the dynamics in the SIR-model, we cannot gain any advantage of equipartition in the SIR-model. However, divide and conquer strategies in this spirit may be interesting when taking into account diffusion terms (see e.g. [14]).

## 4 The importance of the reproduction number and the present situation in Germany

So far it has become evident how essential the (effective) reproduction number is in order to control a pandemic. The importance of this factor can be generalised to models beyond the SIR-model, see e.g. [16] for an elaborate model in which 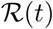 is systematically determined.

We would like to summarise various arguments why 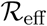 (and 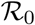 in particular) is the main parameter determining the next months (possibly more) of our lives:

1. The sign of the term 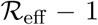 roughly distinguishes between exponential growth or exponential decay of the number of active infections.
2. 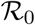 determines the required fraction of the population to reach herd immunity.
3. 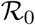 determines the fraction of people who eventually will have contact with the disease if no mitigation measures are imposed.
4. 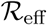 determines the number of *daily new infections*, see e.g. (2.12).
5. The strategy *flattening the curve* requires a fine-tuning of 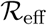 with a precision of < 10%, see Section 2.3. This is why this strategy is disfavoured in this note.
6. The duration *τ* of the strategy *stop the curve* can be estimated by the formula

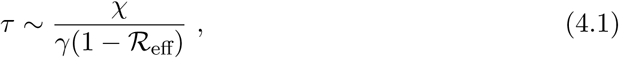

at constant 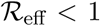 with 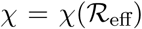 being some number that quantifies how far we are away from being able to track infection chains. At early stages of the pandemic χ is somewhere between 1 < *χ <* 3, also depending on the effective reproduction number.
7. Taking into account diffusion effects one can show that for 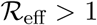 the disease propagates at a speed of [13; 14]

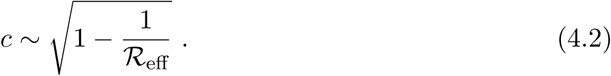 A large reproduction number implies a large diffusion effect. For 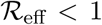 dangerous diffusion effects can be suppressed. Note that diffusion effects may easily trigger a second wave after relaxation of lockdown measures.^2^

In the case of Germany we have 2, 8 · 10^4^ known active infections as of 2020-05-03 [35]. If our goal is to reduce the number of new infections to a few hundred per day one obtains χ ≃ 2 − 3 (as long as the reproduction number is not close to zero), assuming exponential decay of the number of active infections. Keeping 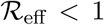 we can therefore estimate the duration of the lockdown to be

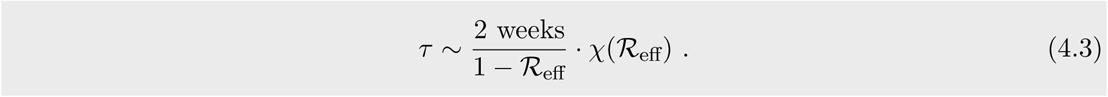

Based on [16] a realistic estimate could be 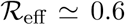 on average during the lockdown. Then we obtain 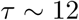 weeks from now. If we were able to achieve and keep 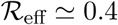 then we need restrictions for *τ* ~ 7 more weeks, and for 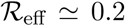, in which *χ ≃* 1.4 we even have *τ* ~ 3-4 weeks. Afterwards, when relaxing the restrictions, it will be crucial to track the origins of every single new infection case, hoping that this tracking helps to keep 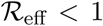 without further lockdowns. It is essential to be well prepared for this highly non-trivial task.

Finally we would like to stress that any strategy following 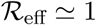 will last for an extremely long time as one can see from (4.3). This is why it is really important to keep the reproduction number significantly below the critical value of 1 in the classical *stop the curve strategy*.

## 5 Conclusion

In this note we have demonstrated the basic mechanics of pandemics using the SIR-model. Despite its simplicity it captures most of the relevant challenges we are facing in the Covid-19 pandemics. Therefore, we expect that most of the results are also implied by more sophisticated models.

In a short review of the SIR-model applied to the Covid-19 pandemics we have seen that the strategy *flattening the curve* suffers from a necessary fine-tuning of the effective reproduction number as well as a precise estimation of the capacity of health care systems. This strategy is therefore risky and problematic.

In the main part of this note we approached a *divide and conquer strategy*. We concluded that the idea to isolate just the high risk group instead of having collective lockdowns fails for two reasons. First, the high risk group is most likely too large to develop herd immunity with respect to the whole population. Second, despite exposing only rather young and healthy people to the virus, the number of daily infections to be expected will probably still exceed Germany’s health care system. Although the result on this strategy is negative, we find the problem of quantifying this scenario interesting in itself, resulting in nice inequalities (3.4) and (3.7) with useful applications in the development of new containment strategies.

Finally, we gave several reasons why the (effective) reproduction number is essential when it comes to control the pandemic. Therefore, a monitoring of this parameter is and remains necessary. This, in turn, requires systematic testing. In the case of Germany we estimated that our present restrictions are presumably necessary for a few more months until we are in the position to track the origins of individual infections. A precise exit strategy involves both the size of the reproduction number and the number of new infections each day. The duration until the exit can then be estimated by (4.3). This formula is not meant to be exact but it provides a rough perspective for the next months. The more we can suppress the reproduction number towards zero the earlier we may be able live with far less restrictions.

## Data Availability

All data used are available via the links provided in the references.

https://github.com/CSSEGISandData/COVID-19

## Acknowledgement

This note has been written out of private interest in this topic. Hence, no funding is to be reported.

## Footnotes

1 Once immunity gets lost the SIR-model can be easily extended to the SIRS-model (see e.g. [12]).

2 This has been pointed out several times by Christian Drosten in his podcast *Das Coronavirus-Update* at NDR.

## Notes

### Competing Interest Statement

The authors have declared no competing interest.

### Funding Statement

This preprint has been written out of private interest in the topic of epidemiology. Hence, no funding is to be reported.

